# In search for the SARS-CoV-2 protection correlate: A head-to-head comparison of two quantitative S1 assays in a group of pre-characterized oligo-/asymptomatic patients

**DOI:** 10.1101/2021.02.19.21252080

**Authors:** Raquel Rubio-Acero, Noemi Castelletti, Volker Fingerle, Laura Olbrich, Abhishek Bakuli, Roman Wölfel, Philipp Girl, Katharina Müller, Simon Jochum, Matthias Strobl, Michael Hoelscher, Andreas Wieser, on behalf of the KoCo19 study team

## Abstract

**Background:** Quantitative serological assays detecting response to SARS-CoV-2 infection are urgently needed to quantify immunity. This study analyzed the performance and correlation of two independent quantitative anti-S1 assays in oligo-/asymptomatic individuals from a previously characterized population-based cohort.

**Methods:** A total of 362 samples included 108 from individuals who had viral RNA detected in pharyngeal swabs, 111 negative controls and 143 samples with positive serology but not confirmed by RT-PCR. Blood plasma was tested with quantitative assays Euroimmun Anti-SARS-CoV-2 QuantiVac ELISA (IgG) (EI-S1-IgG-quant) and Roche Elecsys^®^ Anti-SARS-CoV-2 CoV-2 S (Ro-RBD-Ig-quant), which were compared with each other and with confirmatory tests, including wild-type virus micro-neutralization (NT) and GenScript^®^cPass™. Results were analyzed using square roots *R* of coefficients of determination for association among continuous variables and non-parametric tests for paired comparisons.

**Results:** Quantitative anti-S1 serology correlated well with each other (96%/97% for true-positives and true-negatives, respectively). Antibody titers decreased over time (from <30 days to >240 days after initial positive RT-PCR). Agreement with GenScript-cPass was 96%/99% for true-positives and true-negatives, respectively, for Ro-RBD-Ig-quant and 93%/97% for EI-S1-IgG-quant. Ro-RBD-Ig-quant allowed a distinct separation between positive and negative values, and less non-specific reactivity compared with EI-S1-IgG-quant. Raw values (with 95% CI) ≥28.7 U/mL (22.6–36.4) for Ro-RBD-Ig-quant and ≥49.8 U/mL (43.4–57.1) for EI-S1-IgG-quant predicted virus neutralization >1:5 in 95% of cases.

**Conclusions:** Both quantitative anti-S1 assays, Ro-RBD-Ig-quant and EI-S1-IgG-quant, may replace direct neutralization assays in quantitative measurement of immune protection against SARS-CoV-2 in certain circumstances in the future.

**Key points:** Two quantitative anti-S1 assays showed similar performance and a high level of agreement with direct virus neutralization and surrogate neutralization tests, arguing for their utility in quantifying immune protection against SARS-CoV-2.

## Introduction

Severe acute respiratory syndrome coronavirus 2 (SARS-CoV-2), the causative agent of coronavirus disease 2019 (COVID-19) emerged in Wuhan, China, in December 2019, and within months caused millions of infections and deaths across the globe [1]. Despite multiple interventions, including social distancing, wearing of protective equipment in public and introducing enhanced disinfection procedures, the number of infected individuals worldwide continued to rise beyond the end of 2020 [2].

The gold standard for diagnosis of acute COVID-19 is molecular detection of the viral RNA by reverse transcription-polymerase chain reaction (RT-PCR) [3]. In addition, antibody testing can be used to detect humoral immune responses after the infection. Immunoassays detecting anti-SARS-CoV-2 antibodies can be especially valuable to confirm the extent of population exposure and to quantify vaccine responses [4, 5].

Seroconversion typically starts 5–7 days after SARS-CoV-2 infection [4]. All antibody types (IgA, IgG and IgM) can be detected within the same time frame around week 2–4, and the IgG response persists the longest [4, 6]. The most important targets of humoral response are the nucleocapsid protein (N), involved in viral RNA replication and viral assembly, and parts of the trimeric spike complex, in particular the receptor-binding domain (RBD) of S1 which interacts with the angiotensin-converting enzyme 2 (ACE2) receptor on human cells [7, 8]. Antibodies that bind to RBD in a way that prevents its attachment to the host cell have a convincing functional likelihood to neutralize the virus and are viewed as the key indicator of immune protection [9-11]. In line with these observations, the spike protein became the leading antigen target in vaccine development [12].

Accumulating data suggest that high titers of IgG in convalescent plasma correlate with the presence of neutralizing antibodies, which may correlate with protection against infection [8, 13-15]. However, the long-term persistence of neutralizing antibodies and the degree of protection they confer, as well as the degree and clinical significance of seroconversion in asymptomatic individuals remain largely unknown.

A number of immunoassays from different manufacturers are currently available and have been compared directly in several head-to-head studies [16-19]. Since most are qualitative in nature, the emergence of quantitative assays is needed for a precise evaluation of immune response to viral antigens. Importantly, a reliable quantitative assay can be used to quantify protection in different settings (e.g. mild disease or vaccination). Such evaluation will require robust data on quantitative assay performance and its correlation with available neutralization tests.

Here, we present the results of a direct comparison of two novel quantitative anti-S1/RBD antibody tests applied to a subset of samples derived from a prospective population-based cohort study of COVID-19 incidence/prevalence in Munich, Germany. The ongoing KoCo19 study investigates the prevalence of SARS-CoV-2 infections among a randomly selected cohort, analyzes transmission within households and risk factors, and compares the performance of various immunoassays in testing asymptomatic and oligosymptomatic individuals [20]. The primary results were reported elsewhere [21, 22]. This manuscript reports the analysis of the performance and correlation of quantitative Euroimmun Anti-SARS-CoV-2 QuantiVac ELISA (IgG) assay that recognizes S1 (hereafter called EI-S1-IgG-quant) and quantitative Elecsys^®^ Anti-SARS-CoV-2 S pan-Ig assay that recognizes S1 RBD (hereafter called Ro-RBD-Ig-quant). Both assays were compared with previously described qualitative primary assays, namely Euroimmun Anti-SARS-CoV-2 ELISA IgG (hereafter called EI-S1-IgG) [23, 24] and Elecsys Anti-SARS-CoV-2 Roche anti-N pan-Ig (hereafter called Ro-N-Ig) [25]. The primary tests were also assessed alongside assays that confirm infection, including direct virus neutralization test (NT) with SARS-CoV-2 wild-type virus (SARS-CoV-2 strain MUC-IMB-01 isolated in January 2020), GenScript^®^cPass™ (hereafter called GS-cPass) and Mikrogen-*recom*Line-N/RBD IgG line (hereafter called MG-N and MG-RBD) immunoassays. The WHO reference sera (NIBSC code: 20/268) were measured in replicates (n=3) to standardize the results [26].

## Patients and methods

### Study design and participants

Samples were derived from the population-based prospective COVID-19 cohort KoCo19 from Munich, Germany [20]. Out of the total 6658 samples analyzed previously [22], 362 (due to having NT arrays and all other confirmatory tests) were included in this analysis. These included samples from i. asymptomatic or oligosymptomatic individuals who had at least one RT-PCR positive SARS-CoV-2 test on a pharyngeal swab (true positive samples), ii. those who did not have an RT-PCR positive test, but experienced seroconversion in at least one of the primary tests used (‘other seropositive’ samples) and iii. negative controls – partially blood donors collected before the surge of SARS-CoV-2/COVID-19, and negative samples obtained during the pandemic (true negative samples). All samples were collected during the same time period (between April and June 2020), except for true negative samples from blood donors (October 2019 and March 2020; i.e. before and after the seasonal common cold period). Samples were defined ‘other seropositive’ if a positive result yielded in one of the serological tests, suggesting a likely but unconfirmed SARS-CoV-2 infection. To ensure a dataset with exclusively independent variables, only one serum sample per participant was used for analyses.

The study was approved by the Ethics Commission of the Faculty of Medicine at Ludwig-Maximilians-Universität Munich (20-275-V) and the protocol is available online (www.koco19.de) [20].

### Laboratory assays

Blood samples were obtained as previously described [20]. Briefly, blood was collected in ethylenediaminetetraacetic acid-coated tubes, refrigerated and maintained at 4°C from the moment of extraction until centrifugation to separate the cell pellet from the plasma. Plasma samples were analyzed and stored at –80°C in temperature-controlled biobank freezers; freeze-thaw cycles were minimized to avoid sample degradation.

The presence of antibodies was analyzed using appropriate assay kits according to the manufacturers’ instructions.

An overview of all assays, their cut-off values and readouts are shown in **Table 1**.

**Table 1.**
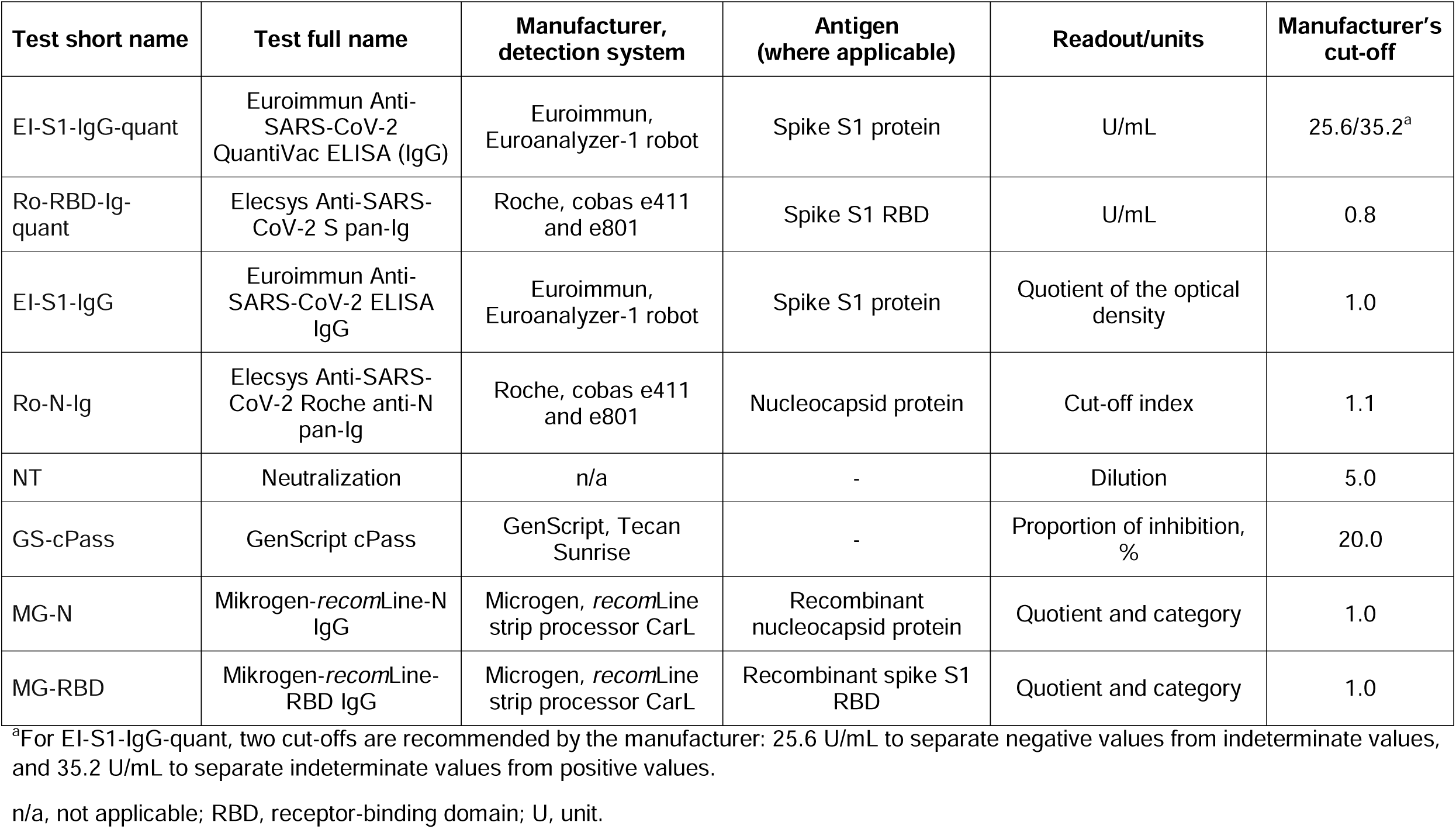
Performance of primary tests and tests confirming infection

### Primary assays

EI-S1-IgG-quant and EI-S1-IgG results were measured on a Euroanalyzer-1 robot (Euroimmun, Lübeck, Germany). For the qualitative assay EI-S1-IgG, presented values show quotients of the optical density measurements given by the manufacturer’s software. For the quantitative assay EI-S1-IgG-quant, values are shown in units per milliliter (U/mL). Two cut-offs were applied as recommended by the manufacturer: 25.6 U/mL to separate negative values from indeterminate values, and 35.2 U/mL to separate indeterminate values from positive values (**Table 1**). Values between 1–120 U/mL represent linear range, samples with values below 1 U/mL were assigned a categorical value of 0, whereas samples with values above 120 U/mL were diluted 1:4 with sample buffer (Euroimmun, Lübeck, Germany) and measured again.

Ro-RBD-Ig-quant and Ro-N-Ig results were measured on cobas e411 and/or e801 modules (Roche, Mannheim, Germany). For the qualitative assay Ro-N-Ig, values correspond to the sample cut-off index. For the quantitative assay Ro-RBD-Ig, values are shown in U/mL. Manufacturer cut-off was applied to separate negative and positive values. Values between 0.4–250 U/mL represent linear range. Samples with values below 0.4 U/mL were assigned a categorical value of 0, whereas samples with values above 250 U/mL were diluted 1:10 with sample diluent buffer (Roche, Mannheim, Germany).

### Assays confirming infection

Confirmatory testing was performed using micro-virus neutralization assays (NT) as described previously [27], with the exception that confluent cells were incubated instead of adding cells following neutralization reaction, and the serum dilutions started with 1:5 instead of 1:10. Samples with a titer <1:5 were classified as NT-negative, and samples with a titer ≥1:5 as NT-positive.

Binding inhibition was measured using the SARS-CoV-2 surrogate virus neutralization test (GS-cPass; GenScript^®^, Piscataway, New Jersey, USA) according to the manufacturer’s instructions. Photometric measurements were performed using the Tecan Sunrise (Tecan, Männedorf, Switzerland). Binding inhibition was calculated in percentages (range from –30% to 100%; cut-off was 20% as recommended by the manufacturer).

The *recom*Line SARS-CoV-2 IgG line immunoassay (MG-N and MG-RBD; Mikrogen, Neuried, Germany) based on nitrocellulose strips with recombinant SARS-CoV-2 antigens N and RBD was measured using the fully automated *recom*Line strip processor CarL (Mikrogen, Neuried, Germany) according to the manufacturer’s instructions. Raw values were presented in arbitrary units, and manufacturer’s cut-offs were applied.

### Statistical analysis

Only one sample per participant was included in the statistical analyses; in case of individuals with multiple blood samples, the sample with the most comprehensive dataset was included. For multiple measurements with complete datasets, only the first measurement was considered; for operational replicates the latest measurement was included. Assay comparison was performed as described previously [22]. Statistical analysis and visualization was performed using software R, version 4.0.2 (https://cloud.r-project.org/). Square roots *R* of coefficients of determination (*R*^*2*^) were calculated for continuous variables. Paired sample comparisons were performed using Wilcoxon-sign-rank tests; multiple group comparisons were performed using Kruskal-Wallis tests, followed by *post hoc* Dunn tests and the Benjamini-Yekutieli adjustment for pairwise comparisons [28].

## Results

A total of 362 samples from the KoCo19 cohort were included in the analysis: 108 samples from individuals who had viral RNA detected in pharyngeal swabs (true positives), 143 ‘other seropositive’ samples and 111 negative controls [20].

### Performance of anti-S1 tests

The diagnostic accuracy measurements of all primary and confirmatory tests are presented in **Table 2**.

**Table 2.**
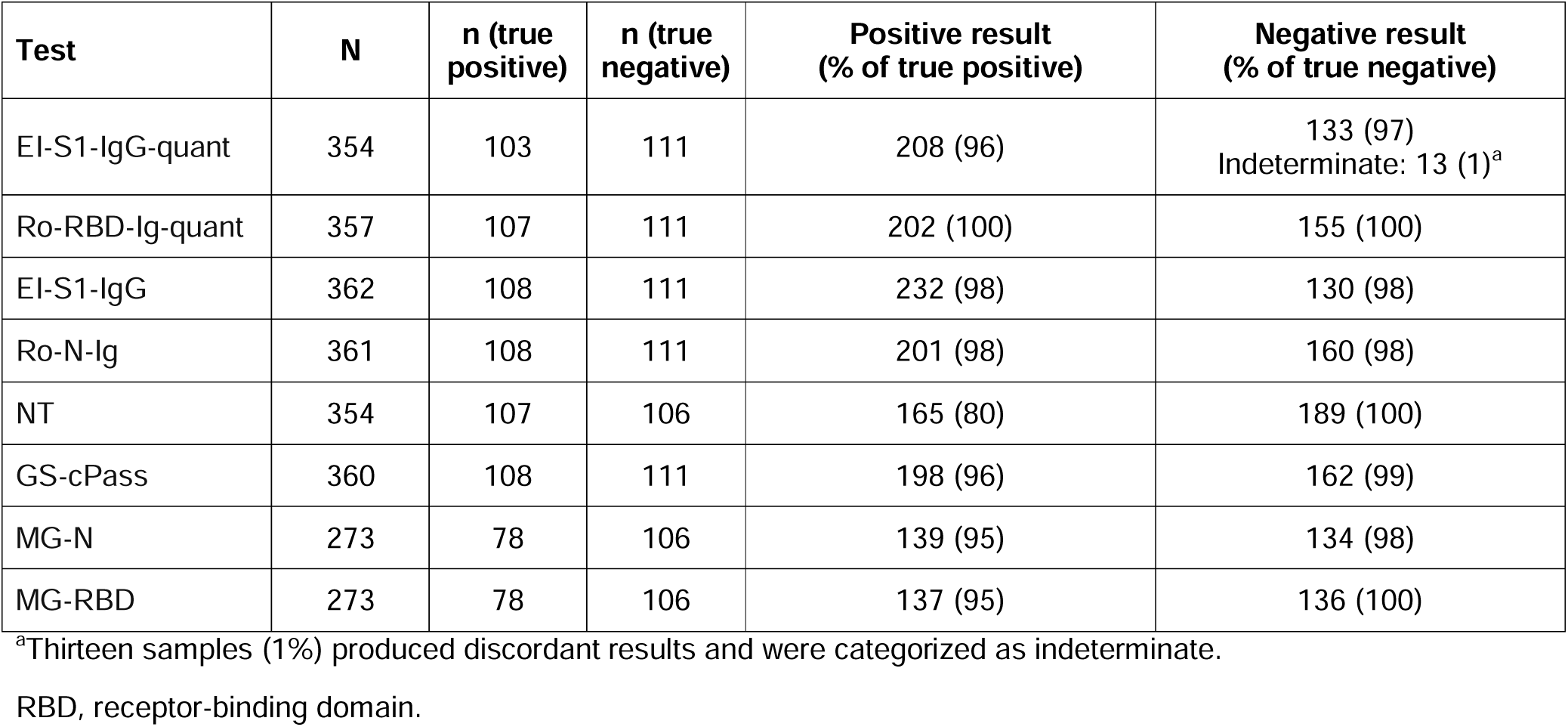
Performance of primary tests and confirmatory tests

The raw value distributions of Ro-RBD-Ig-quant on n=357 evaluable samples and EI-S1-IgG-quant on n=354 evaluable samples are presented in **Figure 1**. Both assays showed a bimodal distribution. Ro-RBD-Ig-quant assay showed a better signal spread with a clear separation of true negative and true positive samples, and did not produce discordant results. Ro-RBD-Ig-quant detected 100% of the positive and 100% of the negative samples, whereas EI-S1-IgG-quant detected 96% of the positive and 97% of the negative samples. Thirteen samples produced discordant results in EI-S1-IgG-quant and were categorized as indeterminate as they did not meet criteria for the positive or negative categories (**Table 2**). Titer values of true positive samples with available data on time between RT-PCR and blood sampling for Ro-RBD-Ig-quant (n=232) and EI-S1-IgG-quant (n=228) are shown in **Figure 2**. Values were widespread in the cohort with <30 days between RT-PCR and antibody test for both assays. The mean titer values tended to decrease over time, with statistically significant differences between value distribution in the cohort with <30 days vs cohort with >240 days for both assays (p<0.0001). After 240 days, the majority (80%) of Ro-RBD-Ig-quant values remained in the positive range whereas almost half of EI-S1-IgG-quant values no longer met the positivity threshold.

**Figure 1.**
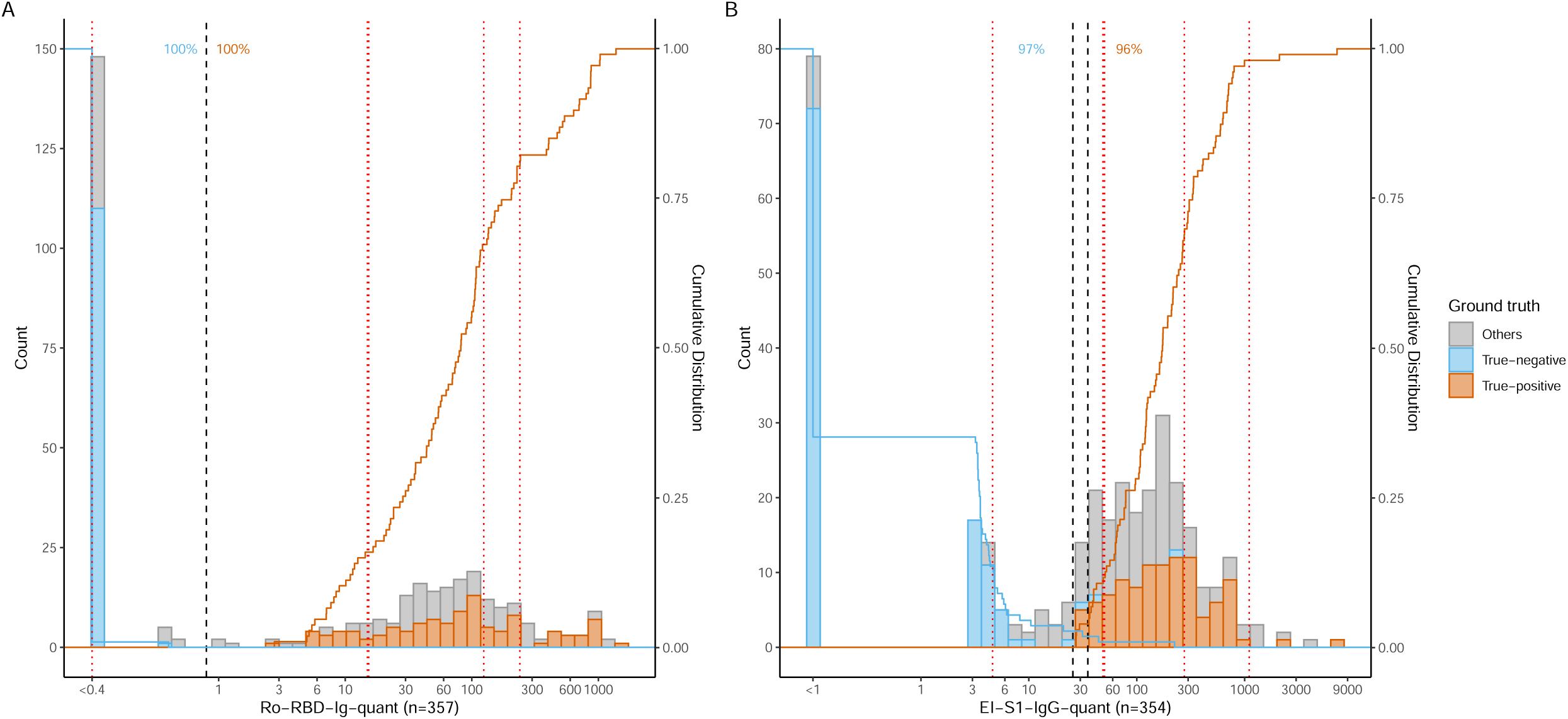
Performance of primary tests Ro-RBD-Ig-quant (A) and EI-S1-IgG-quant (B) Black dashed lines represent manufacturers’ cut-off values and red dotted lines represent WHO standards (from the left to the right: 20/142, 20/144 and 20/140 for panel A (20/140 and 20/144 for panel B) with almost identical values, 20/148, 20/150). Histograms show counts of individual samples, whereas solid blue and orange lines show cumulative distribution of true positive and true negative samples. Orange and blue numbers give the percentage of true positive and true negative samples, which were correctly detected by the tests.

**Figure 2.**
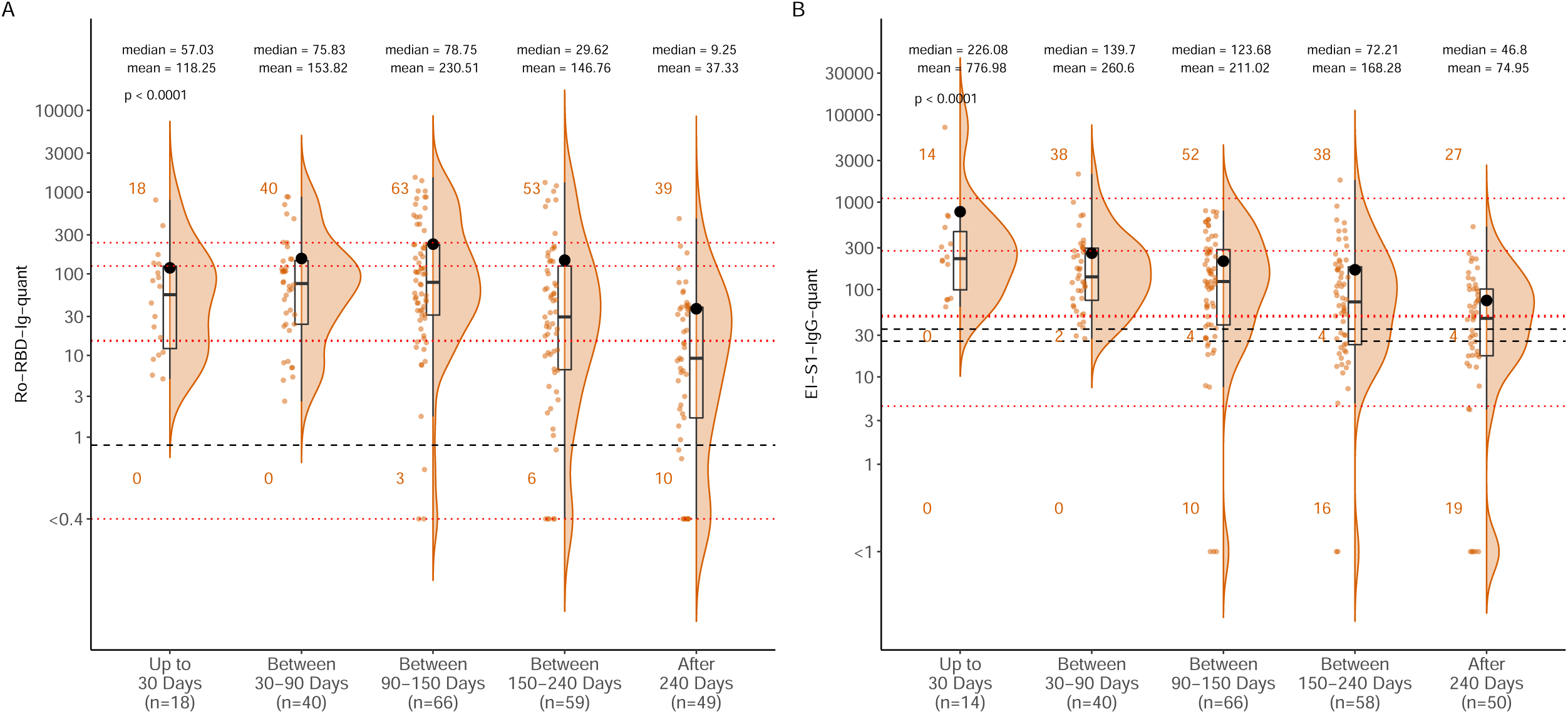
Time-dependent values of positive samples in primary tests Ro-RBD-Ig-quant (A) and EI-S1-IgG-quant (B) Black dashed lines represent manufacturers’ cut-off values and red dotted lines represent the WHO standards (from the bottom to the top: 20/142, 20/144 and 20/140 for panel A (20/140 and 20/144 for panel B) with almost identical values, 20/148, 20/150). Assay results were categorized according to the time after the positive RT-PCR test (<30 days, 30–90 days, 90–150 days, 150–240 days, and >240 days). Plots show the individual read-out (orange dots), a density estimate (orange area), the 25-, 50- and 75-percentiles (black box), and the mean (black dot); mean and median numbers are included for each group. Pairwise comparison between groups after adjustment for multiple comparison are shown in Table S1.

### Concordance between quantitative and semi-quantitative anti-S1 tests

To allow for comparison of scale, results of individual assays with the WHO reference panel (NIBSC 20/168) are presented in **Table 3**.

**Table 3.**
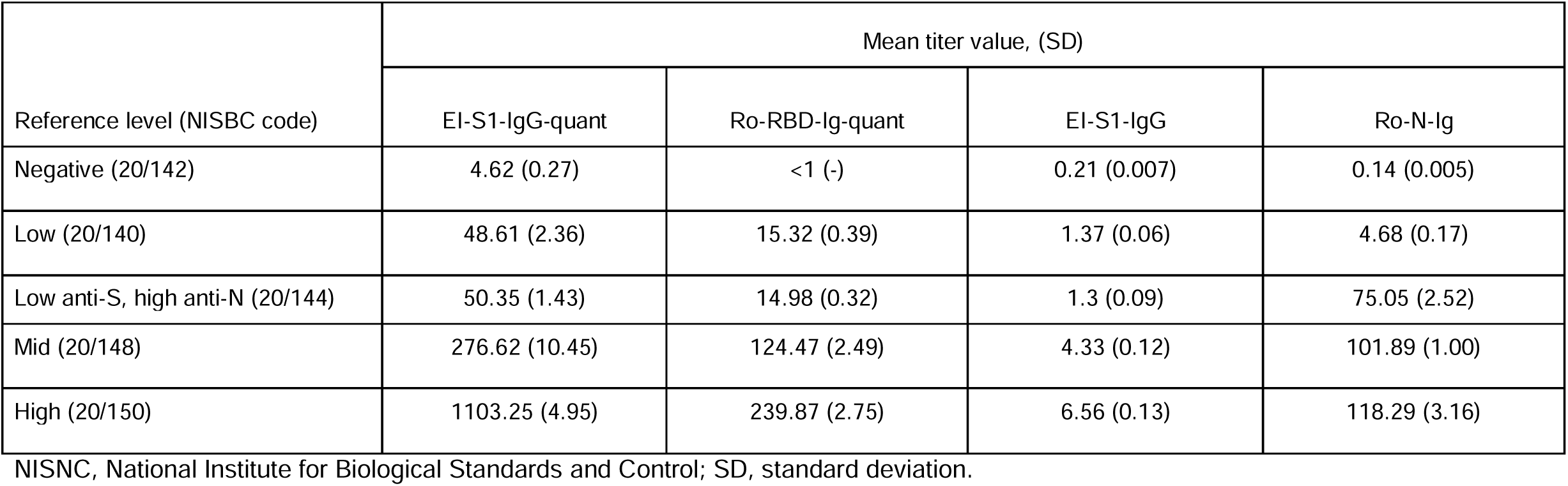
Results from individual assays with the WHO reference panel for anti-SARS-CoV-2 immunoglobulin (NIBSC code: 20/268)

Pairwise comparison of primary tests is shown in **Figure 3**; agreement of 95–98% was observed for all comparisons. When categorical values were excluded, Ro-RBD-Ig-quant showed a high numerical correlation with EI-S1-IgG (*R*=0.72, p<0.0001; **Figure 3A**), while the numerical correlation with Ro-N-Ig was lower (*R*=0.34; p<0.0001; **Figure 3B**). EI-S1-IgG-quant showed a high numerical correlation with EI-S1-IgG (*R*=0.55, p<0.0001; **Figure 3C**) while with Ro-N-Ig the correlation was lower (*R*=0.2, p<0.001; **Figure 3D**). A high level of categorical agreement was observed between Ro-RBD-Ig-quant and EI-S1-IgG-quant with a high correlation (96% of positive samples and 97% of negative samples; *R*=0.5, p<0.0001; **Figure 3E**). Notably, Ro-RBD-Ig-quant gave a clearer separation of positive and negative values than EI-S1-IgG-quant; EI-S1-IgG-quant showed many values at the intermediate range and some non-specific reactivity among the negative samples (3%).

**Figure 3.**
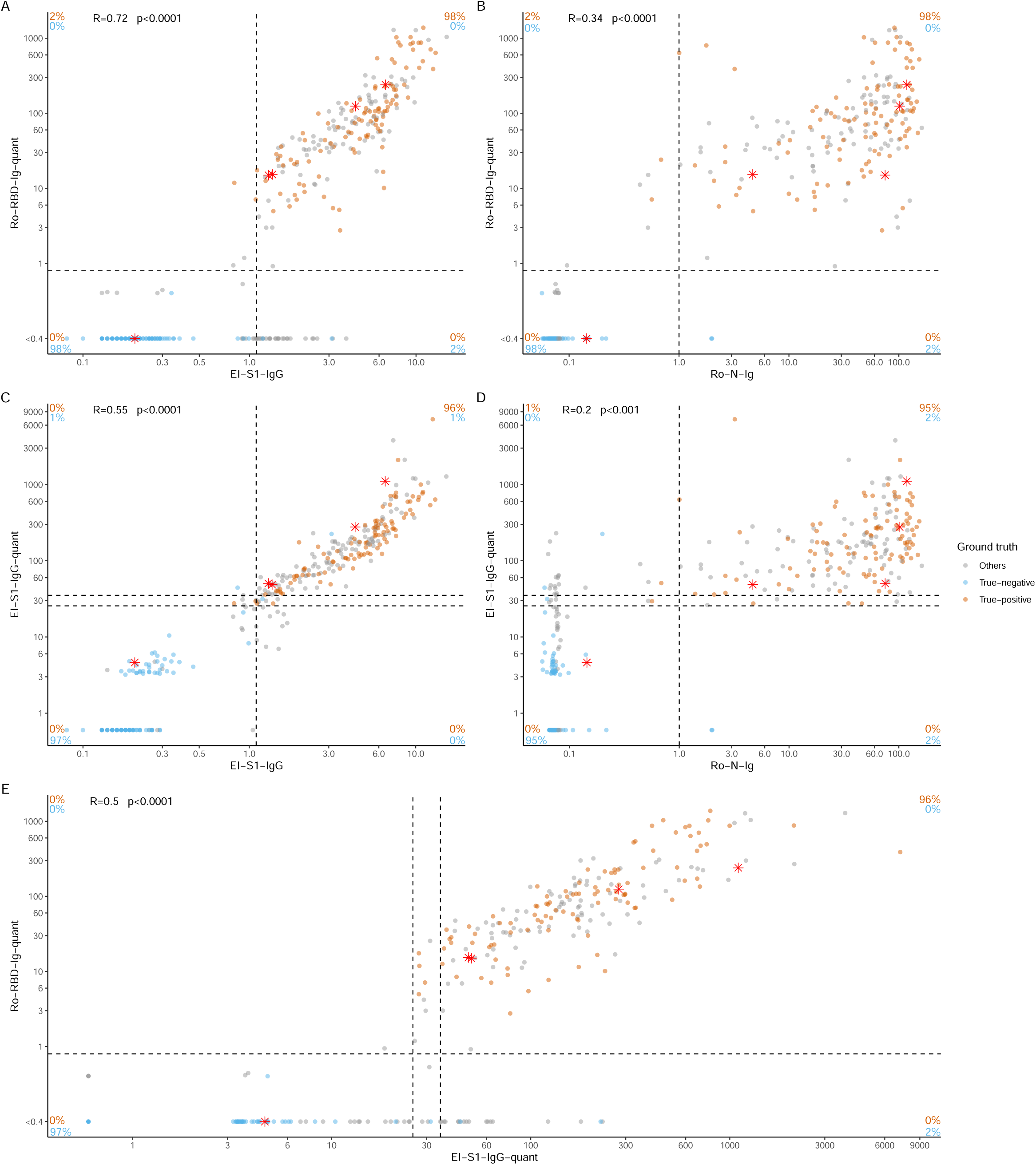
Pairwise comparison of primary tests. Bivariate comparisons shown as scatter plots for quantitative Ro-RBD-Ig-quant vs non-quantitative EI-S1-IgG-quant (**A**) and vs non-quantitative Ro-N-Ig (**B**); for quantitative EI-S1-IgG-quant vs non-quantitative EI-S1-IgG-quant (**C**) and vs non-quantitative Ro-N-Ig (**D**); for quantitative Ro-RBD-Ig-quant vs quantitative EI-S1-IgG-quant (**E**). Dashed lines represent manufacturers’ cut-off values. Red asterisks represent the WHO-standards (from the left to the right: 20/142, 20/144 and 20/140 for panels A and C (20/140 and 20/144 for panels B, D and E) with almost identical values, 20/148, 20/150). Orange and blue numbers give the percentage of true positive and true negative samples, which were correctly detected by the tests. Square root R of coefficients of determination is given for association among continuous variables.

### Concordance with tests confirming infection

Ro-RBD-Ig-quant values showed significant increases between NT dilution categories (p<0.001), with mean values increasing from 39.64 in the NT dilution category <1:5 to 486.24 in the NT dilution category >1:80 (**Figure 4A**). Notably, NT at dilution 1:5 still contained approximately 20% of true positive samples. Ro-RBD-Ig-quant also showed a high categorical agreement and correlation with GS-cPass (96%/99%, *R*=0.54, p<0.0001; **Figure 4B**).

**Figure 4.**
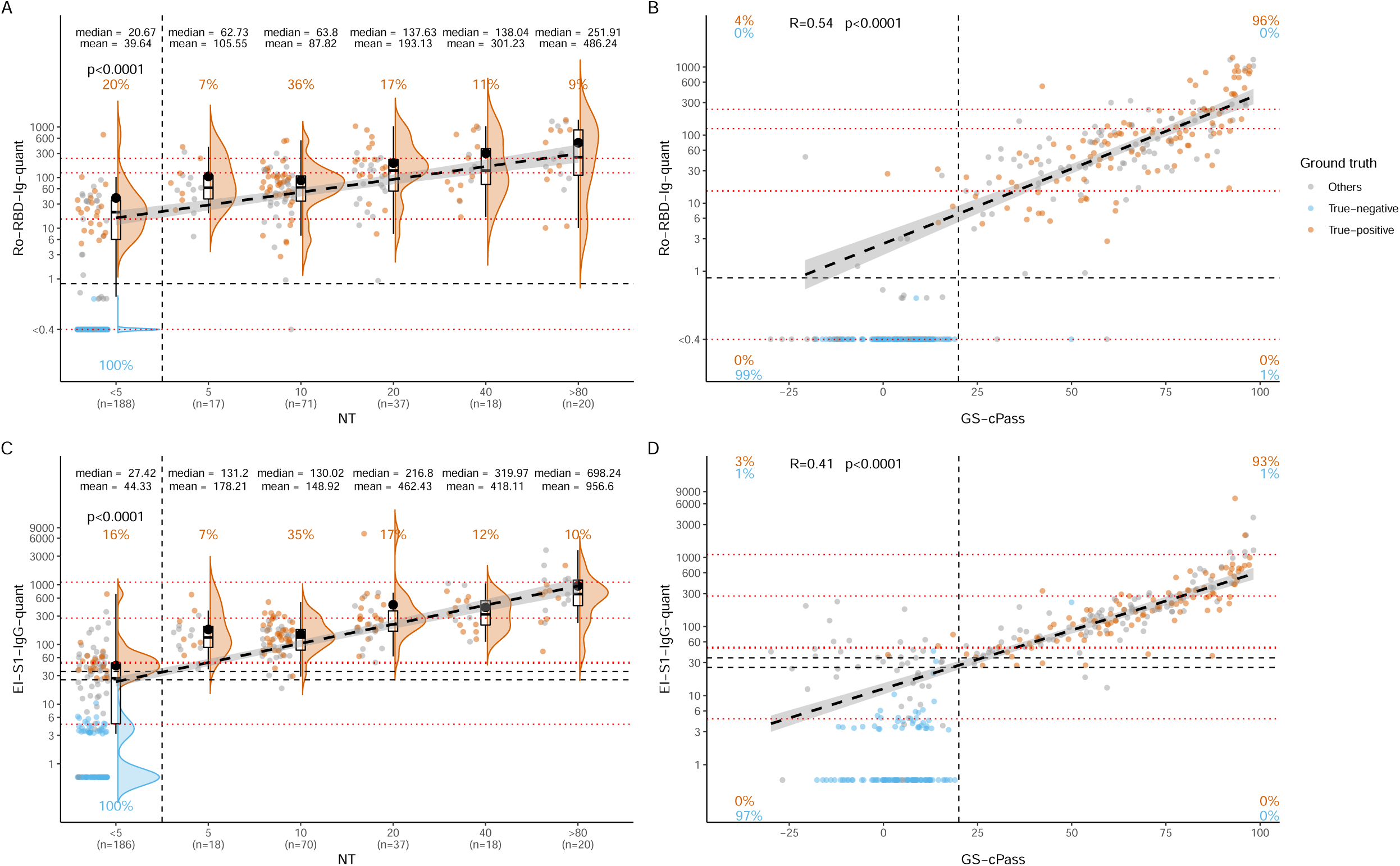
Pairwise comparison of primary tests with confirmatory tests. Bivariate comparisons shown as violin and scatter plots for quantitative Ro-RBD-Ig-quant vs NT at indicated dilutions (**A**) and vs GS-cPass (**B**) and for quantitative EI-S1-IgG-quant vs NT at indicated dilutions (**C**) and vs GS-cPass (**D**). Black dashed lines represent manufacturers’ cut-off values and red dotted lines represent the WHO-standards (from the bottom to the top: 20/142, 20/144 and 20/140 for panel A (20/140 and 20/144 for panel C) with almost identical values, 20/148, 20/150). Orange and blue numbers give the percentage of true positive and true negative samples, which were correctly detected by the tests. Bold dashed lines are linear fit and grey areas surrounding them represent 95% CI; for the interested region, the polynomial fit was within the 95% CI of the linear fit. Square root *R* of coefficients of determination is given for association among continuous variables. Pairwise comparison between NT dilution categories (for **A** and **C**) after adjustment for multiple comparison are shown in Table S2.

EI-S1-IgG-quant values also showed significant increases between NT dilution categories, with mean values of EI-S1-IgG-quant increasing from 44.33 (NT dilution <1:5) to 956.6 (NT dilution >1:80; **Figure 4C**). NT at dilution 1:5 still contained approximately 16% of true positive samples. EI-S1-IgG-quant showed a high level of correlation with GS-cPass (93%/97%, *R*=0.41, p<0.0001; **Figure 4D**), although some unspecific reactivity in the negative samples was detected for EI-S1-IgG (3%).

The predictive value (95% accordance of the positive predictive value) of the two quantitative tests at different thresholds was investigated through the alignment of their results with NT dilution categories ≥1:5 and ≥1:10, and cPass categories ≥20% and ≥30%. The lowest Ro-RBD-Ig-quant and EI-S1-IgG-quant values [with 95% CI] for which cPass is ≥20% (6.99 and 27.49, respectively) or ≥30% (11.60 and 40.62) and NT is ≥1:5 (28.67 and 49.78) or ≥1:10 (51.41 and 104.06; **Figure 5**). These values refer to the intersection of the linear fit with the selected values for cPass and NT of **Figure 4**.

**Figure 5.**
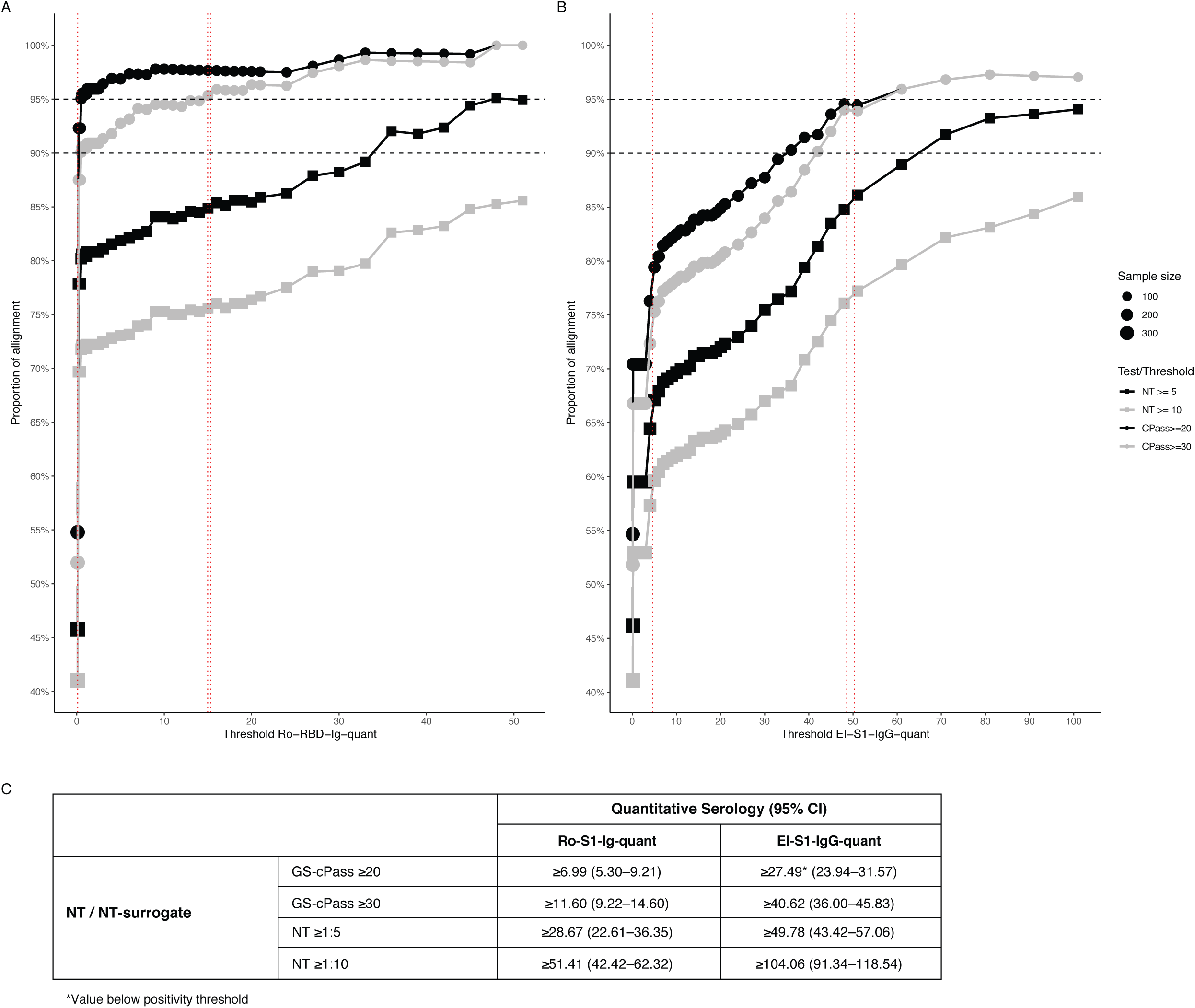
Predictive value of quantitative tests. Shown as proportion of alignment of Ro-RBD-Ig-quant (**A**) and EI-S1-IgG-quant (**B**) with NT dilution categories ≥1:5 and ≥1:10 and cPass categories ≥20% and ≥30%. Red dotted lines represent the WHO-standards (from the left to the right: 20/142, 20/144 and 20/140 for panel A [20/140 and 20/144 for panel B] with almost identical values). Table (**C**) details the lowest values (95% accordance of the positive predictive value [95% CI]) of the quantitative tests for which NT is ≥1:5 and ≥1:10 and cPass is ≥20% and ≥30%. CI, confidence interval; NT, neutralization

Ro-S1-Ig-quant showed a high level of correlation with line blot assay MG-RBD and a lower numerical correlation with MG-N (*R*=0.44, p<0.0001 and *R*=0.32, p<0.001, respectively; **Supplementary Figure 1A,B**). EI-S1-IgG-quant showed a high level of correlation with MG-RBD (*R*=0.46, p<0.0001; **Supplementary Figure 1C**), but the agreement with MG-N was not statistically significant (*R*=0.15, p=0.089; **Supplementary Figure 1D**).

## Discussion

Since the surge of the COVID-19 pandemic, the number of serological assays continues to increase, and the comparative assessment of their analytical performance is essential to inform strategies in diagnostic, epidemiological and vaccination studies. In this study, we performed a head-to-head direct comparison of two independent quantitative assays directed against S1 and compared their performance with two qualitative primary assays and several assays that confirm infection, including direct virus neutralization.

The quantitative tests Ro-RBD-Ig-quant and EI-S1-IgG-quant showed a high level of correlation when used in a population cohort containing samples from mostly oligo- or asymptomatic individuals; both assays showed categorical agreement with Ro-N-Ig, micro-virus neutralization assay, GS-cPass and *recom*Line. This suggests both tests can detect correlates of neutralization, which is understood to mediate humoral protection following SARS-CoV-2 infection. While the mean titers for both assays tended to decrease after their peak (∼ 1 month or ∼ 3 months after infection for EI-S1-IgG-quant and Ro-RBD-Ig-quant, respectively) to >240 days after positive RT-PCR, a higher proportion of Ro-RBD-Ig-quant values remained positive after 240 days.

Finally, both quantitative assays showed a good level of concordance with each other, with Ro-RBD-Ig-quant performing slightly better in terms of clearer separation of positive and negative samples and less non-specific reactivity.

Currently, the most reliable method of detecting antibody response indicative of protection is direct virus neutralization; however, this test requires intact virus and has to be performed under biosafety level 3 conditions, making it infeasible for large scale studies and diagnostic routine testing [17]. There are also numerous different protocols for direct viral neutralization with poor overall comparability. Commercial high-throughput tests use parts of viral proteins instead and combine these with other components of chemiluminescence detection or enzyme-linked immunosorbent assay (ELISA) [3, 6]. Although all viral proteins are likely to elicit some degree of immune response, most efforts concentrated on measuring antibodies directed against N and S1/RBD so far [5]. Some studies suggest that S1 may be the optimal antigen for SARS-CoV-2 serological tests, as it is more sensitive than RBD and more specific than S trimer [29]; however, this assumption could not be confirmed in this study. Quantitative anti-S1 assays will be a valuable tool for measuring antibody responses to SARS-CoV-2. Importantly, quantitative assays will allow us to precisely enumerate and compare antibody titers in individuals who had severe disease, mild disease, asymptomatic individuals and those who achieved immunity after vaccination. The assays may also be applied to screen for plasma samples that contain specific high-affinity neutralizing antibodies and help identify potential donors of plasma for convalescent plasma therapy [30]. Once established and rigorously validated, these assays may replace the current gold standard of direct neutralization, which requires handling at biosafety level 3 and has severe limitations in signal resolution at the lower end of the range.

In this study, both EI-S1-IgG-quant and Ro-RBD-Ig-quant showed a high level of correlation with direct virus micro-neutralization and surrogate neutralization test, GS-cPass. For example, raw values above 28.7 U/mL for Ro-RBD-Ig-quant and above 49.8 U/mL for EI-S1-IgG-quant, respectively, predicted virus neutralization >1:5 in 95% of cases. We may hypothesize that when the value of the quantitative tests is above the predictive value (e.g. 95%), there is little benefit in performing NT and that this could act as a surrogate marker for neutralizing titers e.g., after mass vaccinations or post-infection.

Our results suggest that both quantitative assays may be useful in future studies aimed to assess immunization efficiency, determine the degree of herd immunity and estimate how long the response persists over time.

## Data Availability

Data are subject to data protection regulations and can be made available upon reasonable request to the corresponding author. To facilitate reproducibility and reuse, the code used to perform the analyses and generate the figures was made available on GitHub (https://github.com/koco19/lab_epi) and has been uploaded to ZENODO (http://doi.org/10.5281/zenodo.4300922, DOI 10.5281/zenodo.4300922) for long-term storage.

## Acknowledgements

The authors thank all study participants for their trust, time, data, and samples, and all personnel at study centers and in the field for their contributions. ELECSYS is a trademark of Roche. All other product names and trademarks are the property of their respective owners.

## Author contributions

AW, MH, RRA designed the study, RRA, VF, RW, PG, KM performed laboratory analysis, MH, RW, LO, SJ, MS, RRA and AW assisted with analysis of the data. NC did data curation, data preparation and data cleaning, and together with AB, performed statistical analysis and figure generation. LO and AW provided samples and selected the patients from the KoCo19 study. SJ, MS, NC, RRA, and AW prepared the manuscript. All authors have read and approved the final version of the manuscript.

