## Supplementary figures and images for "In search for the SARS-CoV-2 protection correlate: A head-to-head comparison of two quantitative S1 assays in a group of pre-characterized oligo-/asymptomatic patients"

### Supplemental Figure 1

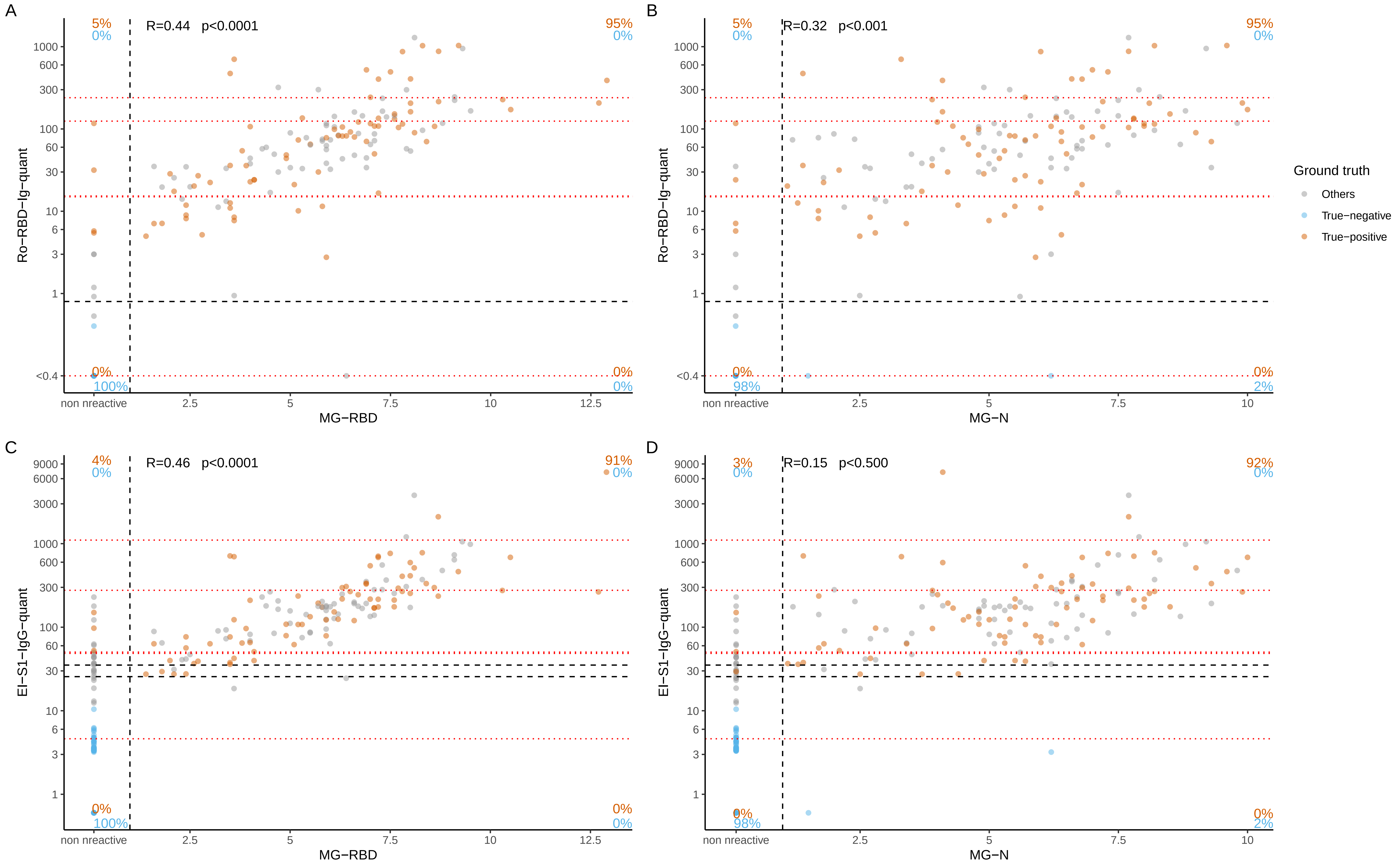
