## Supplemental Material for "In search for the SARS-CoV-2 protection correlate: A head-to-head comparison of two quantitative S1 assays in a group of pre-characterized oligo-/asymptomatic patients"

**Supplementary material**

**Supplementary Tables**

**Table S1.** Pairwise comparison between the time-dependent groups after adjusting for multiple comparison

| **Group comparison** | **Adjusted p value** | |
| --- | --- | --- |
|  | **Ro-RBD-Ig-quant** | **EI-S1-IgG-quant** |
| Up to 30 days - Between 30-90 days | 1.000 | 0.71 |
| Up to 30 days - Between 90-150 days | 1.000 | 0.28 |
| Up to 30 days - Between 150-240 days | 1.000 | <0.05* |
| Up to 30 days - After 240 days | <0.05* | <0.001** |
| Between 30-90 days - Between 90-150 days | 1.000 | 1.000 |
| Between 30-90 days - Between 150-240 days | 0.169 | 0.06 |
| Between 30-90 days - After 240 days | <0.001** | <0.001** |
| Between 90-150 days - Between 150–240 days | <0.05* | 0.16 |
| Between 90-150 days - After 240 days | <0.0001*** | <0.01* |
| Between 150-240 days - After 240 days | <0.05* | 0.30 |

*Significant difference

**Table S2. Pairwise comparison between the NT dilution categories after adjusting for multiple comparison**

|  | **Adjusted p-v** | |
| --- | --- | --- |
| **Group comparison** | **Ro-RBD-Ig-quant** | **EI-S1-IgG-quant** |
| <5 - 5 | <0.01* | <0.0001*** |
| <5 - 10 | <0.001** | <0.0001*** |
| <5 - 20 | <0.0001*** | <0.0001*** |
| <5 - 40 | <0.0001*** | <0.0001*** |
| <5 - >80 | <0.0001*** | <0.0001*** |
| 5 - 10 | 1.000 | 1.000 |
| 5 - 20 | 0.861 | 0.448 |
| 5 - 40 | 0.380 | 0.142 |
| 5 - >80 | 0.058 | <0.01* |
| 10 - 20 | 0.084 | <0.05· |
| 10 - 40 | 0.052 | <0.05· |
| 10 - >80 | <0.01* | <0.0001*** |
| 20 - 40 | 1.000 | 1.000 |
| 20 - >80 | 0.388 | 0.145 |
| 40 - >80 | 1.000 | 1.000 |

*Significant

**Supplementary Figure Legends**

**Supplementary Figure 1. Pairwise comparison of primary tests with line blot tests**

Bivariate comparisons shown as scatter plots for quantitative Ro-RBD-Ig-quant vs MG-RBD (**A**) and vs MG-N (**B**) and for quantitative EI-S1-IgG-quant vs MG-RBD (**C**) and vs MG-N (**D**). Black dashed lines represent manufacturers’ cut-off values and red dotted lines represent the WHO-standards (from the bottom to the top: 20/142, 20/144 and 20/140 for panels A and B (20/140 and 20/144 for panel C and D) with almost identical values, 20/148, 20/150). Orange and blue numbers give the percentage of true positive and true negative samples, which were correctly detected by the tests.
